# Protocol for the Work And Vocational advicE (WAVE) randomised controlled trial testing the addition of vocational advice to usual primary care (Clinical Trials: NCT04543097)

**DOI:** 10.1101/2024.09.11.24313466

**Authors:** G Wynne-Jones, M Lewis, G Sowden, I Madan, K Walker-Bone, CA Chew-Graham, K Bromley, S Jowett, V Parsons, G Mansell, K Cooke, SA Lawton, B Saunders, J Pemberton, C Cooper, NE Foster

## Abstract

**Objectives:** To investigate the effectiveness of adding a brief vocational advice intervention to usual care in reducing the number of days absent from work over a period of 6 months in adults given a fit note by their general practice.

**Design:** Multicentre, pragmatic, two parallel-arm, randomised controlled trial with health economic analyses and nested qualitative study. A computer-generated stratified block randomisation (ratio 1:1) was used to allocate arms.

**Setting:** Participants will be recruited from general practices in the UK.

**Participants:** 720 adults consulting in general practice, for any health condition, and receiving a fit note who have been absent from work for more than two-weeks but less than six months.

**Interventions:** Participants in the intervention arm will be offered usual care and vocational advice delivered by a Vocational Support Worker (VSW) remotely via phone or videoconferencing. Participants in the control arm will be offered usual care.

**Main outcome measure:** Number of days off work over 6 months. Follow-up data collection is via questionnaires at 6 weeks and 6 months.

**Conclusions:** This paper presents the rationale, design and methods of the Work And Vocational advicE (WAVE) trial. The results of this trial will provide evidence to inform primary care practice and guide the development of services to provide support for patients with work absence.

***Trial registration:*** Clinical Trials: NCT04543097

***Protocol number***: Version 5.1

## Background

### Limitations of current occupational healthcare

In the United Kingdom (UK), vocational advice is variable and is often accessible only to those working for larger organisations (1). Therefore, the first port of call for most people who are struggling with their health and work is primary care, where a patient can request a ‘fit note’ to submit to their employer to allow absence from work. The fit note was introduced in 2010 (replacing the ‘sick note’) and encourages clinicians to discuss with a patient whether they are ‘not fit for work’ or ‘may be fit for some work’ with some adaptations (such as phased return to work; altered hours; amended duties; workplace adaptations), with free text space for the clinician to add details. Legislation introduced in 2022 allowed a broader range of clinicians to complete fit notes, including nurses, physiotherapists, occupational therapists, and pharmacists who can now certify a period of work absence and provide advice about working despite health conditions (2). In practice, however, most fit notes certify patients as not fit for work with just 6% using the ‘may be fit’ for work option (3). Whilst evidence is that work is generally good for health, healthcare practitioners report that they struggle to have conversations about work due to lack of confidence, lack of clear guidance around what they should be saying, and because provision of vocational advice is not embedded in general practice (4).

Improving the provision of vocational advice and support in primary care could benefit patients’ health, quality of life, and society through more active participation in the workforce (5). This is pertinent when the number of days lost from work is considered; in 2022 an estimated 185.6 million days were lost due to sickness absence in the UK (6), where approximately £72 million is spent on health-related sickness benefits per year (7). The key drivers of these costs are common illnesses such as musculoskeletal (MSK) disorders, and mental health (MH) conditions (6). Data from NHS Digital show that there is regional variation in fit notes issued across England with the northwest reporting the highest rate per 100,000 patients and London the lowest rate (8). Of concern though is that 42.6% of fit notes are issued for 5 weeks or longer (8): sickness absence is considered long-term after 4 weeks (9), and evidence shows that the longer a person is absent from the workplace the harder it is for them to return (4).

The inclusion of early work-directed interventions has been demonstrated to be effective and cost-effective for conditions such as depression and MSK pain (10–13). Where models of integrated health and occupational advice have been tested, they have led to fewer days of work absence, earlier return-to-work (RTW) and reductions in healthcare use (14–16). Several UK studies have tested interventions to manage work absence in those with health conditions, and their results have informed the WAVE trial (10,17–19).

Learning from our previous Study of Work And Pain (SWAP)(10) randomised trial and nested qualitative research with patients, vocational advisors and General Practitioners (GPs) highlighted that those with at least two weeks of work absence appeared to benefit more from the vocational advice intervention than those with shorter periods of work absence, a finding supported by other research (10,20–23). The SWAP trial offered a vocational advice intervention to patients consulting with MSK pain, and it is not known whether and how the intervention might be amended for use with a broader range of primary care patients, particularly those with common MH conditions. Parallels can be drawn between RTW processes in MSK pain and MH, and the following interventions have been shown to be effective for both types of condition: case management (24); provision of work accommodations (25); addressing obstacles to work that are clinical, psychosocial and organisational (akin to the Flags model) (26); stepped care (27); programmes tailored to individual patients (28); and telephone-delivered interventions (29). Our previous SWAP trial intervention for adults with MSK pain included these evidence-based interventions (10).

Offering vocational support early, in primary care, where most patients with health conditions are seeking healthcare, offers a key potential solution to the current lack of universal provision of vocational advice. The WAVE trial will test whether our previously developed vocational advice intervention can be adapted for all adults consulting in primary care whose health condition is impacting their ability to work.

This protocol is reported in-line with the Standard Protocol Items: Recommendations for Interventional trials (SPIRIT) reporting guidelines (30).

### Aim

The overall aims of the WAVE trial are to determine, in adults consulting in general practice who receive a fit note, whether the addition of a brief vocational advice intervention to usual care leads to fewer days lost from work, and whether offering this vocational advice intervention is cost-effective.

#### Objectives

1. Primary objective To investigate the effectiveness of adding a brief vocational advice intervention to usual care in reducing the number of days absent from work over a period of 6 months in adults who receive a fit note from their general practice.
2. Secondary objectives
  a. Determine the cost-effectiveness of offering the vocational advice intervention in addition to usual care,
  b. Investigate time to RTW and compare this between trial arms,
  c. Investigate factors mediating observed differences in outcomes between the trial arms (e.g. RTW self-efficacy, health symptoms and fear avoidance beliefs),
  d. For the nested qualitative interviews: To explore and understand the perspectives and experiences of trial participants, Vocational Support Workers, primary care clinicians, and employers/line-managers about decision-making around work absence and RTW. Understand participants’ experiences of receiving, and the acceptability of, the vocational advice intervention and its delivery in practice.

### Ethical approval

Ethical approval was granted by National Research Ethics Service (NRES) Committee West of Scotland Research Ethics Committee (REC) 5, September 2020 (REC reference: 20/WS/0127).

## Methods

### Trial design

The WAVE trial is a multi-centre, pragmatic, two-parallel arm, randomised controlled trial with a health economic evaluation and nested qualitative study.

### Setting

This trial will take place within the primary care setting in England. General practices in the West Midlands, London and Wessex areas will participate in recruiting participants. The intervention will be delivered remotely by trained VSWs using phone or videoconferencing, with the option of face-to-face delivery.

### Participant eligibility

*Inclusion criteria*

- Adults aged 18 years and over,
- Currently in paid employment (full or part time),
- Current absence from work of at least two consecutive weeks but not more than six continuous months,
- Received a fit note,
- Access to a mobile phone that can receive and respond to SMS text messages,
- Able to read and write English,
- Able to give full informed consent,
- Willing to participate.

*Exclusion criteria*

- Long-term work absence defined as over six continuous months,
- Pregnancy or maternity leave,
- Patients presenting with signs or symptoms indicative of serious illness requiring urgent medical attention (‘red’ flags),
- Severe mental health problems (e.g. severe depression with risk of self-harm, exacerbation of schizophrenia or bipolar disorder, cognitive impairment, or lack of capacity), high vulnerability (e.g. palliative stages of illness, recent bereavement, dementia).

### Participant identification

Potential participants will be identified when they consult at one of the participating general practices and are issued a fit note for time off work. General practices will choose one of three methods to identify and invite potential participants. Figure 1 provides a flow chart of how participants progress through recruitment.

**Figure 1:**
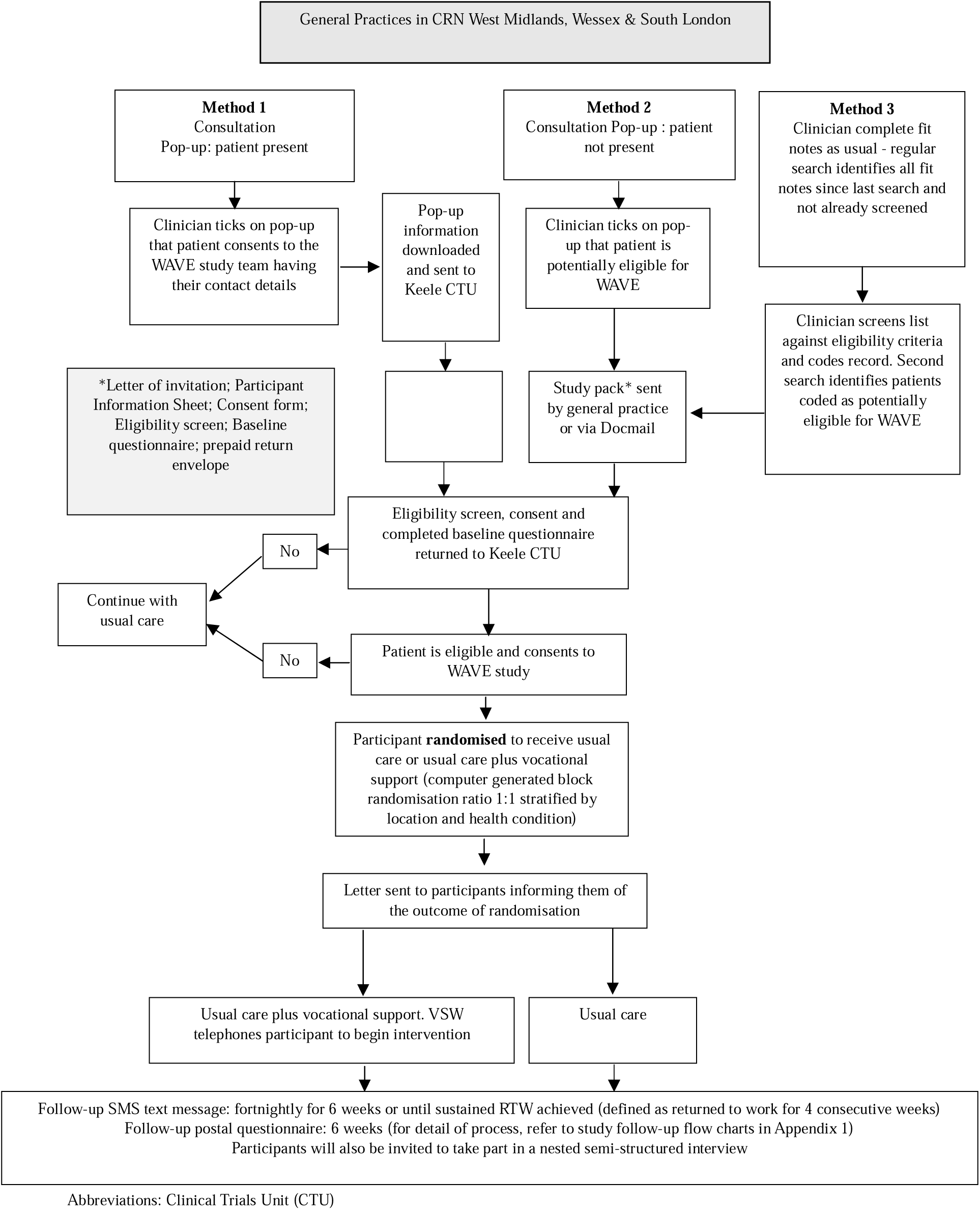
**Participant recruitment**

#### 1. Identification through an automated EMR IT Protocol during “real time” consultations

Identification of potentially eligible trial participants by an automated electronic medical record (EMR) protocol (a “pop-up”) activated when a clinician completes an electronic fit note (eMED3) in the EMR. The specially designed study specific pop-up will only trigger if the patient is aged 18 or over, and there are no clinical codes in a patient’s EMR that match the exclusion criteria. The pop-up will serve several purposes: to flag potentially eligible participants to the consulting clinician; to prompt the clinician to check the patient’s eligibility for the trial by reviewing the list of eligibility criteria (confirmation of eligibility will be automated where possible so that the pop-up does not ‘fire’ for those patients who are clearly ineligible); to prompt the clinician to mention the trial to potentially eligible participants and to ask the patient if they are willing to receive further information about the trial and give their consent to share their contact details with the research team. The clinician will be prompted to provide responses to these items on the pop-up system, and the pop-up will then record, using clinical codes, patients’ eligibility status and consent for further contact.

#### 2. Identification through searches of the general practice EMR after consultation where a pop-up is ‘fired’ on completion of a fit note

Since consultation styles vary, it is possible that some eligible participants will be missed using the above approach, for example if: the clinician does not code the fit note in the EMR until after the patient has left the consultation room; the clinician does not have time to discuss the trial and gain consent for further contact;, the fit note was requested using an online query system so that there was no discussion between the clinician and patient prior to the issue of the fit note. For these situations, a modified pop-up will activate upon entry of the fit note code into the EMR. The modified pop-up will include everything except patient consent to share contact details and the clinician will still have to screen the patient for eligibility and enter eligibility information into the EMR. Tagged records will be downloaded regularly by general practice staff, who will then send a study pack and a letter of invitation on practice headed paper to eligible patients.

#### 3. Identification through back-dated searches of the general practice EMR

To reduce the interruption to consultations by pop-ups and where the patient does not have any direct contact with the clinician, a third method of patient identification may be used. The clinician will issue the fit note as usual, which will trigger a background EMR protocol to automatically screen the patient’s EMR for eligibility. A regular search will be run to identify those patients issued a fit note since the last search. Those patients identified as having been issued a fit note and not meeting any of the exclusion criteria will either (a) be sent a study pack from the practice or, (b) be sent an invitation letter on practice headed paper, containing a link to an electronic consent to contact form so that patients can identify themselves as eligible and consent to being sent a study pack in the post. Where practices use text messaging, an invitation may be sent by text message containing a link to the online consent to contact form.

Practices will have the option to review the list of potentially eligible patients prior to the invitation being sent. Following invitations, the practice will securely transfer a list of invited patients’ NHS numbers, age, and gender to the research team to facilitate accurate communication with the practice about patient participation.

### Participant recruitment

Figure 1 details the participant identification and recruitment processes. Where patients have been sent a study pack, they will be invited to answer:

- whether they are still absent from work (Yes / No),
- whether the duration of their absence is more than 2 weeks (Yes / No),
- whether their absence is less than 6 months (Yes / No),
- whether the fit note is for their employer (Yes / No) (to exclude those issued a fit note for employment support allowance).

Patients who respond ‘No’ to any of these questions will be advised not to complete the remainder of the questionnaire. Participants who respond “Yes” to all the questions will be asked to complete the consent form and baseline questionnaire and return these to Keele Clinical Trials Unit (CTU).

### Consent

The invitation letter will introduce the study and explain, or remind them, how they were selected to be invited. The Participant Information Sheet (PIS) will summarise the study and tell the patient what is involved should they wish to participate. The contact details of Keele CTU will be provided should potential participants have any further questions about the study or have any difficulty in completing the consent form or baseline questionnaire.

Those who wish to participate will be asked to complete, sign and date a consent form confirming that: they have read and understood the PIS and are willing to take part in the study; understand that a questionnaire will be sent at 6 weeks; consent to receive a fortnightly SMS text message to collect data on RTW for up to 6 months; that they understand and consent to randomisation; and that they are aware they can withdraw at any time without giving a reason and that if they do withdraw that their clinical care will not be affected.

Participants may also consent to the optional aspects of the trial, again signing and dating the consent form to confirm that they: may be invited to participate in an interview; understand that their GP will be notified about their participation in the study; were asked to consent to a pseudonymised electronic copy of relevant sections of their general practice medical records to allow authorised members of the research team to extract information relevant to the study.

The returned consent form will be checked for completion. Recruitment will be complete when the returned questionnaire confirms the patient is eligible, they have signed and dated their consent form and completed the primary outcome data. Any missing data from the consent form, eligibility questions or primary outcome, will be followed up by post, telephone or email from Keele CTU.

### Randomisation and allocation concealment

On confirmation of eligibility and receipt of a consent form participants will be randomised by Keele CTU to either usual care or usual care plus the vocational advice by computer-generated stratified block randomisation (ratio 1:1). Stratification will be by centre (West Midlands, London, Wessex) and main health condition resulting in time off work (MH, MSK pain or ‘other’ health condition). All participants will be mailed a letter informing them whether they have been randomised to either the usual care or to usual care plus the vocational advice intervention. A letter will be sent to the GP in participating practices of all randomised participants informing them of their patient’s participation and allocation.

Participants, their treating clinicians, and VSWs cannot be blinded to allocation due to the nature of the intervention. Keele CTU staff who may need to contact participants for minimum data collection by telephone will be blinded to allocation. The data will be analysed independently by two statisticians, one of whom will be blinded to intervention allocation; the other statistician will be unblinded to allow intervention delivery details and content of case report forms (CRFs) to be reported to the TSC / Data Monitoring Committee (DMC) as required.

### Description of intervention and control

#### Control arm

Patients randomised to usual care will be offered usual care, which for most patients, will not include formal vocational advice. Information on usual care received will be collected through participant questionnaires (to include questions about other vocational advice or occupational health services received) and through review of general practice EMR.

#### Intervention arm

Participants who are randomised to the intervention will be offered the vocational advice intervention in addition to usual care. The intervention will be a modification of that developed and successfully delivered in the SWAP trial (10), comprising a stepped care intervention based on the principles of case management (31). A full description of the content, delivery and training associated with the intervention will be published separately. Figure 2 presents a summary of each step of the intervention.

**Figure 2:**
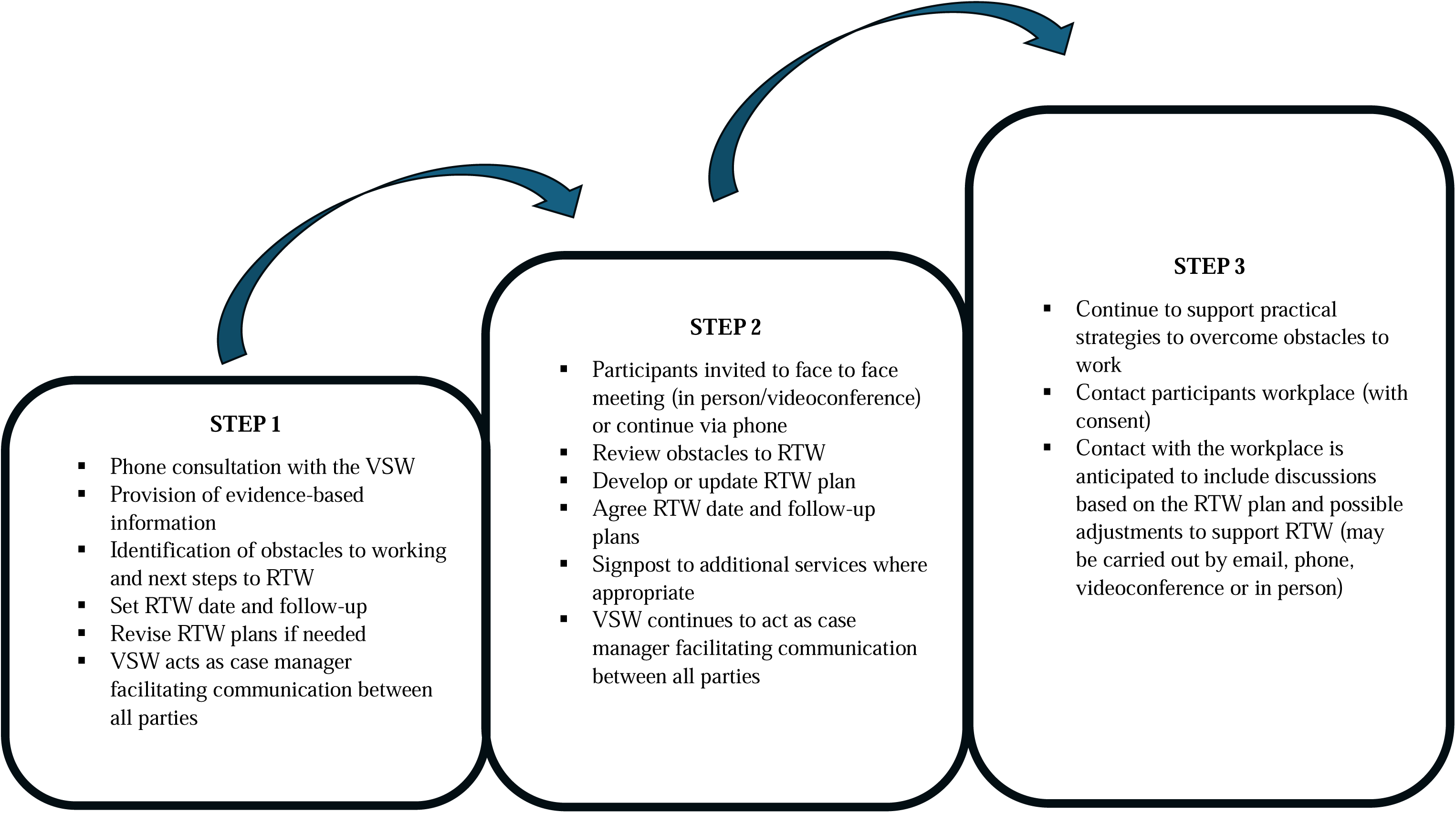
**Overview of the intervention**

### Sample size

The trial is powered to detect a 25% reduction in days off work over 6 months between the intervention and control arms, equating to an Incidence Rate Ratio (IRR) of 0.75 (e.g., mean days off work reduced from 30 days in the control arm to 22.5 days in the intervention arm). A sample size of 720 gives 80-90% power to detect an IRR of 0.75 based on a 5% two-tailed significance test and assumed dispersion parameter of 1.4 (derived from the previous SWAP trial) allowing for 20% loss to follow-up (10).

## Data collection

To collect baseline and outcome data, participants will be sent postal questionnaires shortly after participants receive their fit note, then at 6 weeks and 6 months following randomisation. The baseline questionnaire will include the primary and secondary outcome measures, key anticipated moderators of treatment effect, health economic variables, and demographic information. The questionnaire at 6 weeks will include the primary outcome and key anticipated mediator variables only. The questionnaire at 6 months will be slightly longer and include the primary and secondary outcome measures, and questions about self-reported healthcare use including about any other vocational advice received through employers, health services or any other agency. Table 1 provides a summary of the questionnaire measures and timing of the data collection.

**Table 1:**
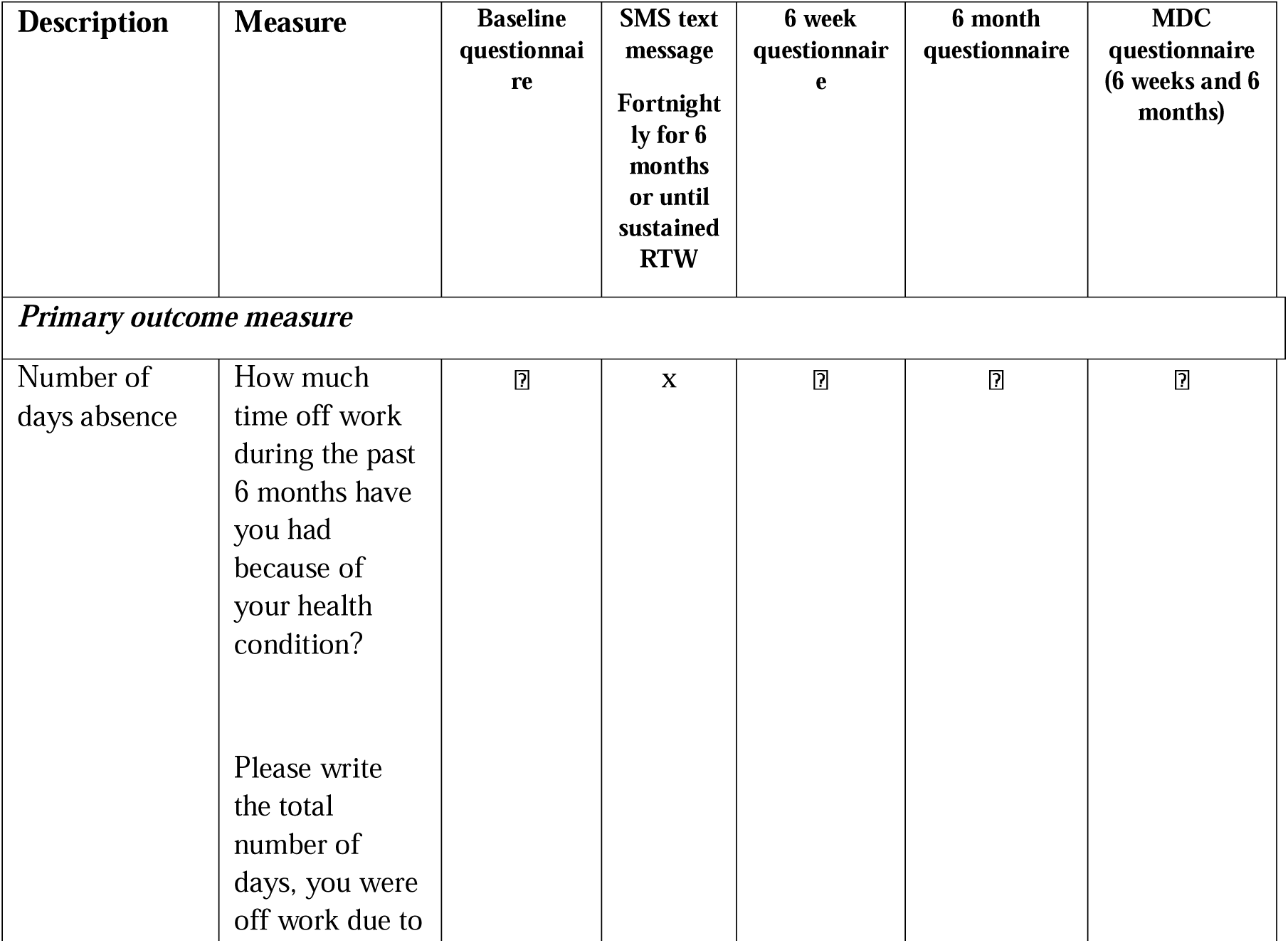
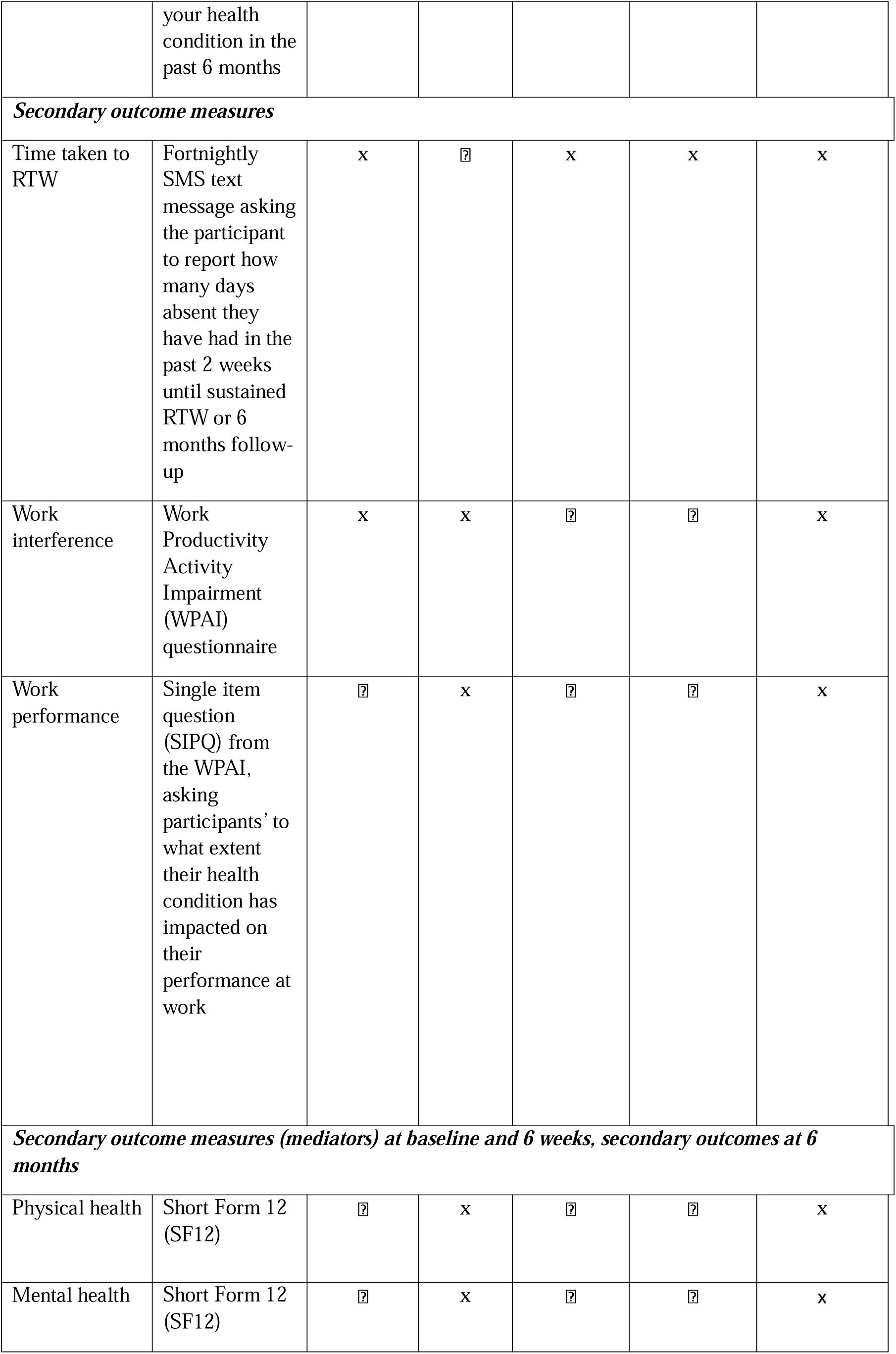
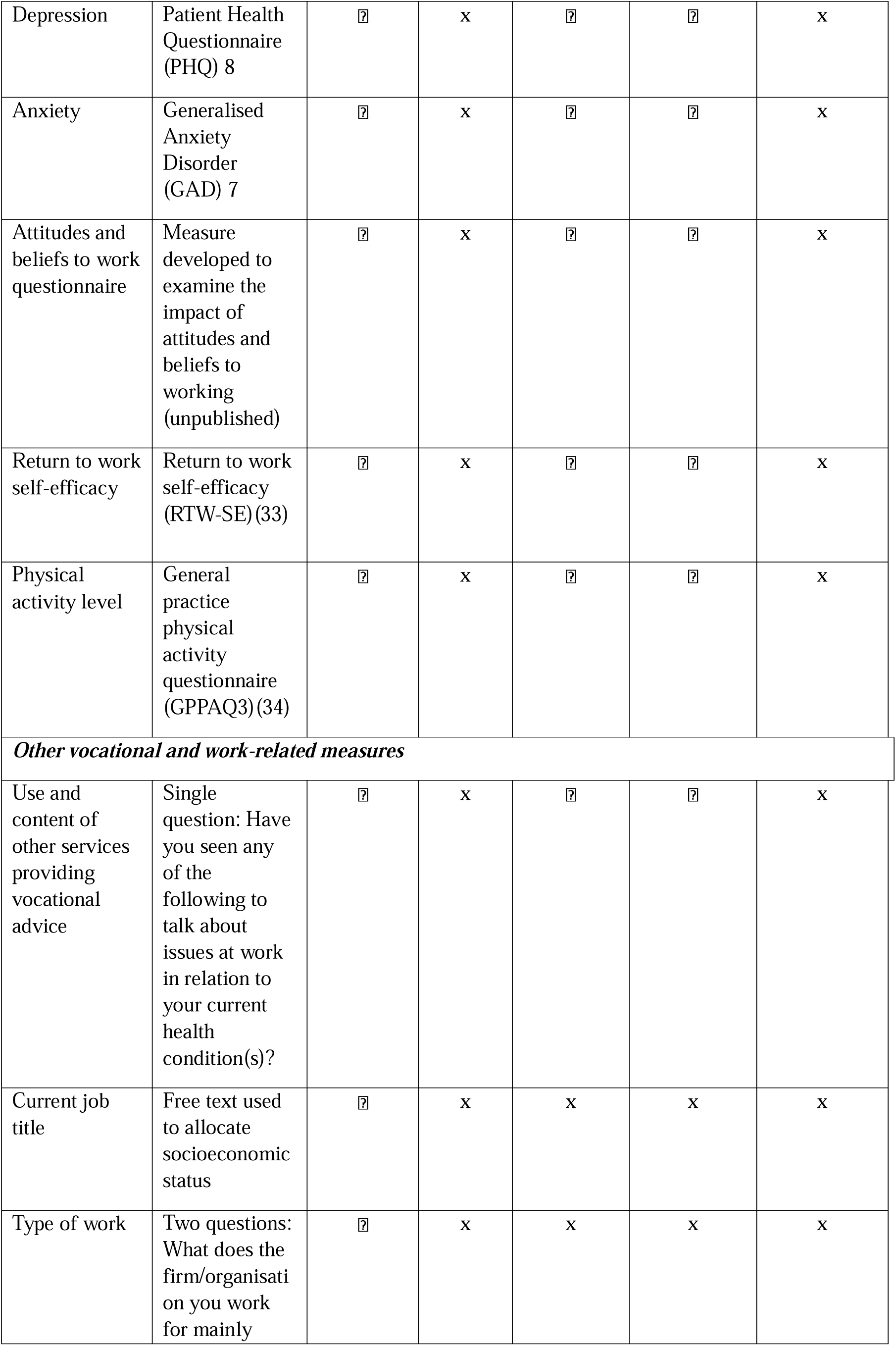
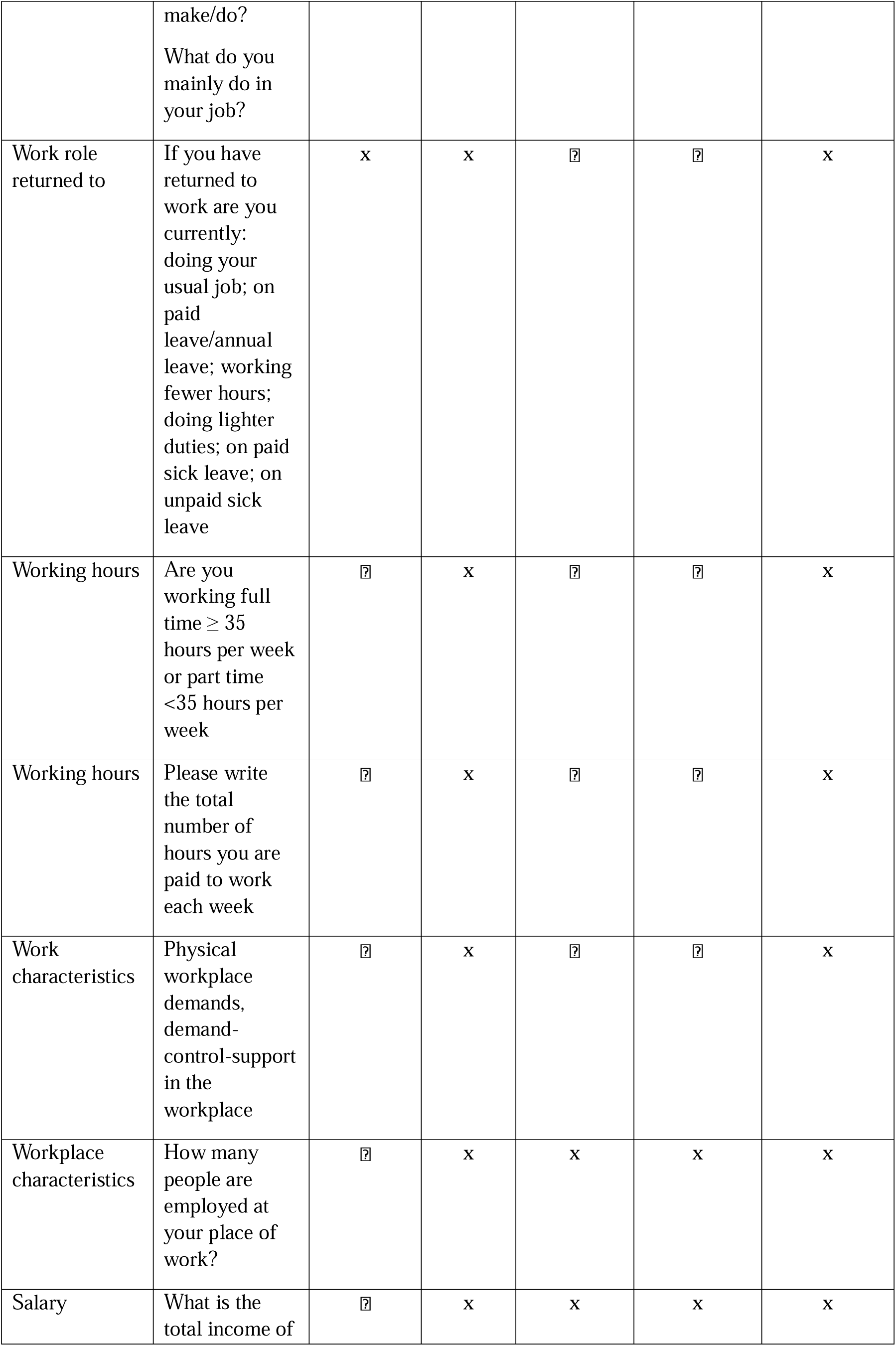
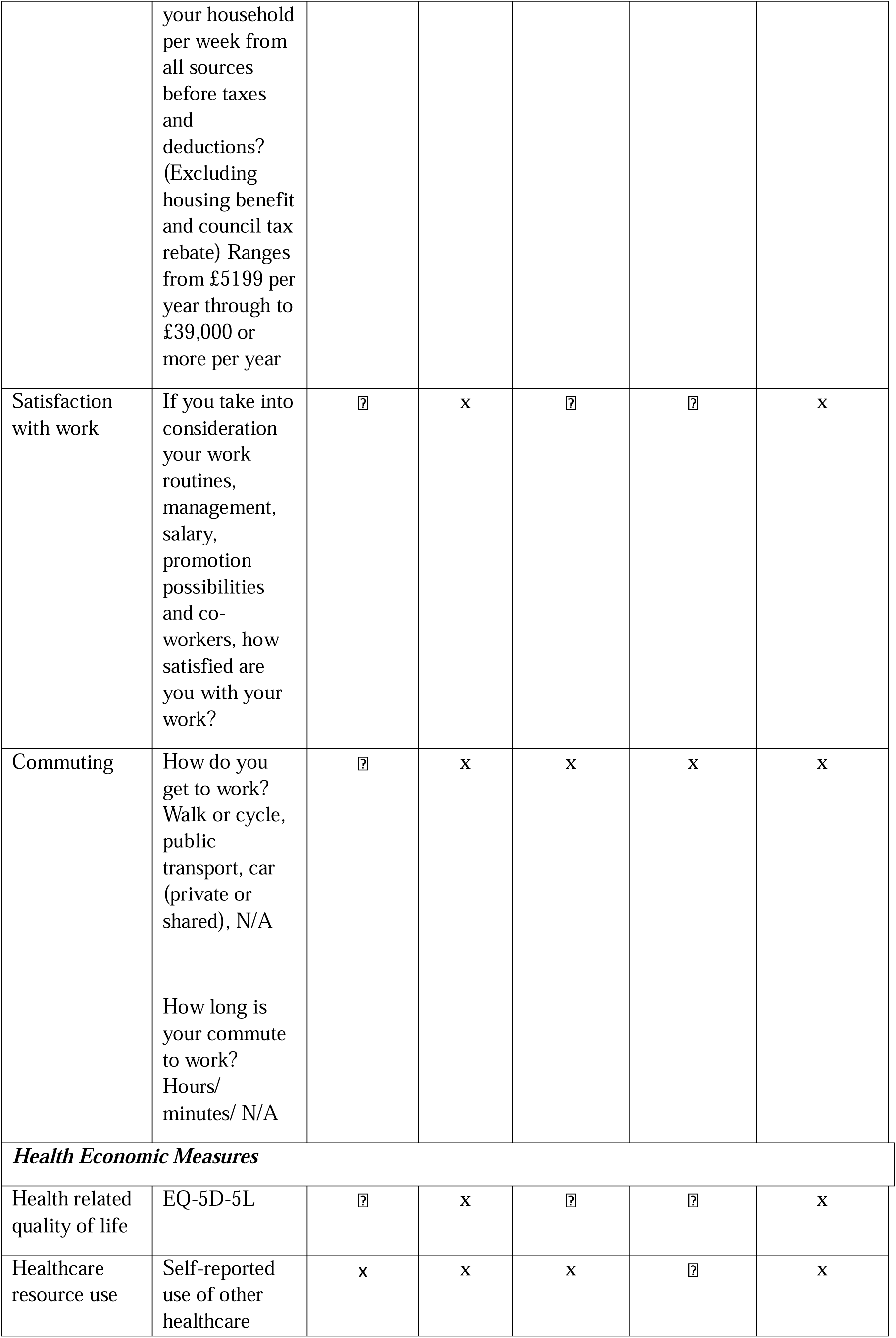
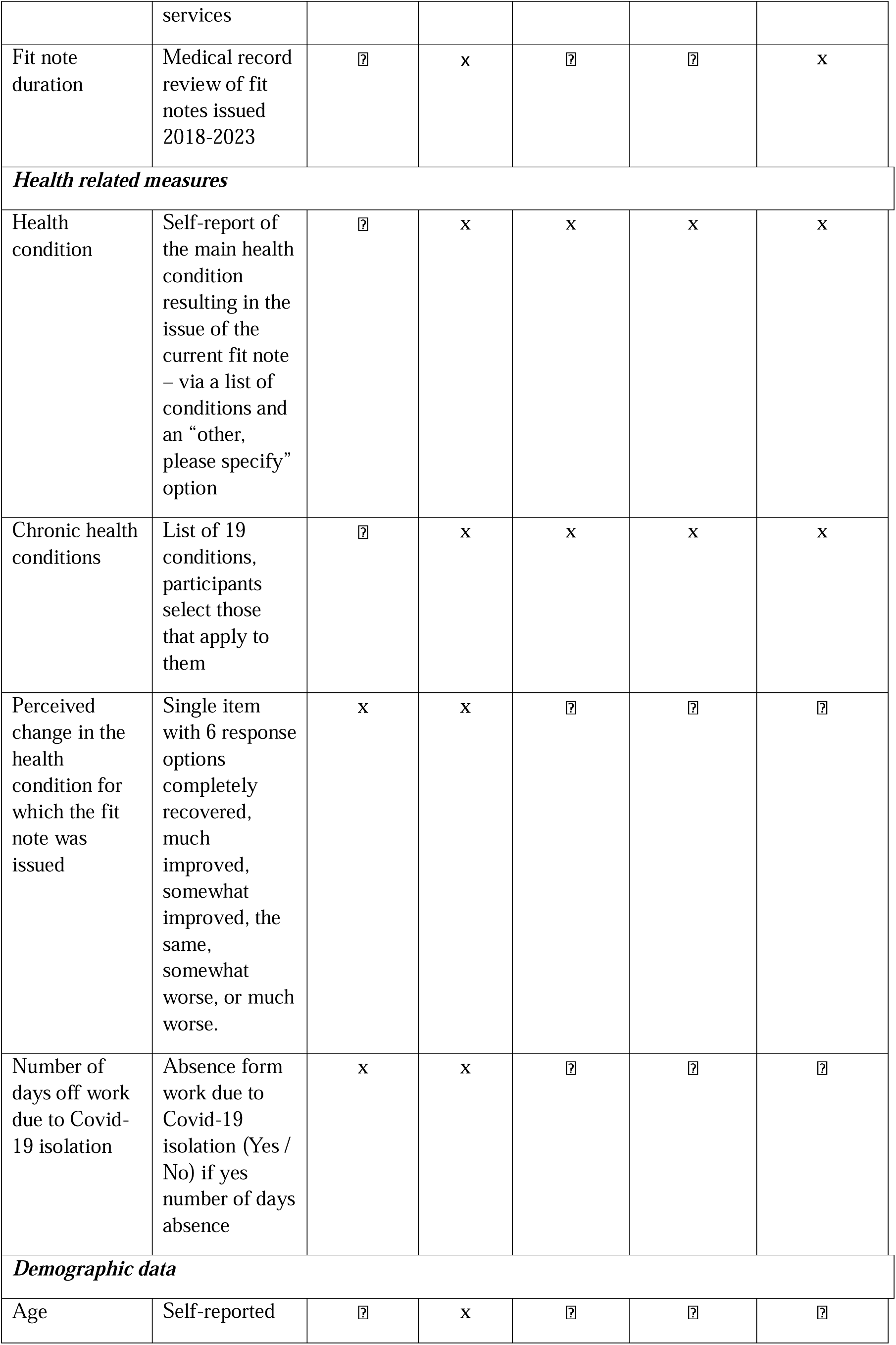
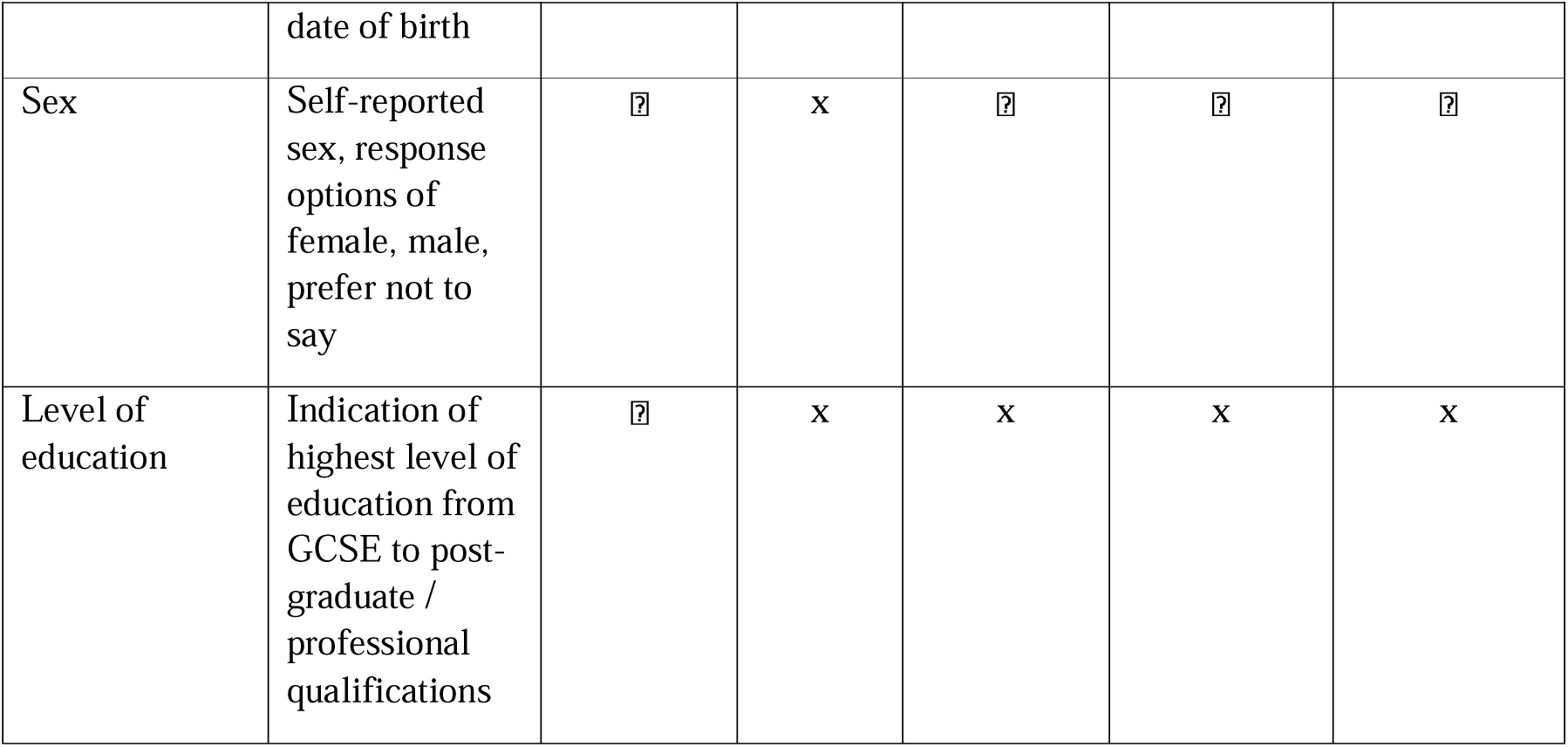
Questionnaire measures and time-points.

| **Description** | **Measure** | **Baseline questionnaire** | **SMS text message**<br>**Fortnightly for 6 months or until sustained RTW** | **6 week questionnaire** | **6 month questionnaire** | **MDC questionnaire (6 weeks and 6 months)** |
| --- | --- | --- | --- | --- | --- | --- |
| ***Primary outcome measure*** |  |  |  |  |  |  |
| Number of days absence | How much time off work during the past 6 months have you had because of your health condition?<br>Please write the total number of days, you were off work due to your health condition in the past 6 months | ? | x | ? | ? | ? |
| **Secondary outcome measures** |  |  |  |  |  |  |
| Time taken to RTW | Fortnightly SMS text message asking the participant to report how many days absent they have had in the past 2 weeks until sustained RTW or 6 months follow-up | x | ? | x | x | x |
| Work interference | Work Productivity Activity Impairment (WPAI) questionnaire | x | x | ? | ? | x |
| Work performance | Single item question (SIPQ) from the WPAI, asking participants' to what extent their health condition has impacted on their performance at work | ? | x | ? | ? | x |
| **Secondary outcome measures (mediators) at baseline and 6 weeks, secondary outcomes at 6 months** |  |  |  |  |  |  |
| Physical health | Short Form 12 (SF12) | ? | x | ? | ? | x |
| Mental health | Short Form 12 (SF12) | ? | x | ? | ? | x |
| Depression | Patient Health Questionnaire (PHQ) 8 | ? | x | ? | ? | x |
| Anxiety | Generalised Anxiety Disorder (GAD) 7 | ? | x | ? | ? | x |
| Attitudes and beliefs to work questionnaire | Measure developed to examine the impact of attitudes and beliefs to working (unpublished) | ? | x | ? | ? | x |
| Return to work self-efficacy | Return to work self-efficacy (RTW-SE)(33) | ? | x | ? | ? | x |
| Physical activity level | General practice physical activity questionnaire (GPPAQ3)(34) | ? | x | ? | ? | x |
| ***Other vocational and work-related measures*** |  |  |  |  |  |  |
| Use and content of other services providing vocational advice | Single question: Have you seen any of the following to talk about issues at work in relation to your current health condition(s)? | ? | x | ? | ? | x |
| Current job title | Free text used to allocate socioeconomic status | ? | x | x | x | x |
| Type of work | Two questions: What does the firm/organisation you work for mainly make/do?<br>What do you mainly do in your job? | ? | x | x | x | x |
| Work role returned to | If you have returned to work are you currently:<br>doing your usual job; on paid leave/annual leave; working fewer hours; doing lighter duties; on paid sick leave; on unpaid sick leave | x | x | ? | ? | x |
| Working hours | Are you working full time $\geq 35$ hours per week or part time $<35$ hours per week | ? | x | ? | ? | x |
| Working hours | Please write the total number of hours you are paid to work each week | ? | x | ? | ? | x |
| Work characteristics | Physical workplace demands, demand-control-support in the workplace | ? | x | ? | ? | x |
| Workplace characteristics | How many people are employed at your place of work? | ? | x | x | x | x |
| Salary | What is the total income of your household per week from all sources before taxes and deductions? (Excluding housing benefit and council tax rebate) Ranges from £5199 per year through to £39,000 or more per year | ? | x | x | x | x |
| Satisfaction with work | If you take into consideration your work routines, management, salary, promotion possibilities and co-workers, how satisfied are you with your work? | ? | x | ? | ? | x |
| Commuting | How do you get to work?<br>Walk or cycle, public transport, car (private or shared), N/A<br>How long is your commute to work?<br>Hours/ minutes/ N/A | ? | x | x | x | x |
| **Health Economic Measures** |  |  |  |  |  |  |
| Health related quality of life | EQ-5D-5L | ? | x | ? | ? | x |
| Healthcare resource use | Self-reported use of other healthcare services | x | x | x | ? | x |
| Fit note duration | Medical record review of fit notes issued 2018-2023 | ? | x | ? | ? | x |
| **Health related measures** |  |  |  |  |  |  |
| Health condition | Self-report of the main health condition resulting in the issue of the current fit note – via a list of conditions and an “other, please specify” option | ? | x | x | x | x |
| Chronic health conditions | List of 19 conditions, participants select those that apply to them | ? | x | x | x | x |
| Perceived change in the health condition for which the fit note was issued | Single item with 6 response options completely recovered, much improved, somewhat improved, the same, somewhat worse, or much worse. | x | x | ? | ? | ? |
| Number of days off work due to Covid-19 isolation | Absence form work due to Covid-19 isolation (Yes / No) if yes number of days absence | x | x | ? | ? | ? |
| **Demographic data** |  |  |  |  |  |  |
| Age | Self-reported date of birth | ? | x | ? | ? | ? |
| Sex | Self-reported sex, response options of female, male, prefer not to say | ? | x | ? | ? | ? |
| Level of education | Indication of highest level of education from GCSE to post-graduate / professional qualifications | ? | x | x | x | x |

### Primary outcome measure

The primary outcome measure is number of days off work over 6 months and will be assessed by asking participants to report how much time off work they have had due to their health condition. See Table 1.

### Secondary outcome measures

Participants will be asked every 2 weeks whether they have returned to work, for a period of 6 months or until a sustained RTW is achieved (defined as return to *any work* for at least 4 weeks). RTW will be measured via SMS text message using the following questions;

- lr Have you returned to work? Yes/No,
- lr If yes, on which date did you return to work e.g. 13SEP2021.

Work interference will be measured using the Work Productivity Activity Impairment (WPAI) questionnaire (32). The WPAI measures impairments to work and activities in the past 7 days and has been validated in many health conditions including MH, MSK pain, respiratory, digestive cardiovascular and other conditions. A single item work productivity question from the WPAI, asking participants to what extent their health condition has impacted their performance at work will also be used as a secondary outcome measure.

Where mediators are collected at 6 months they will also be treated as secondary outcome measures.

### Moderators and mediators

The questionnaires will include anticipated mediators that have been included within the initial logic model underpinning the intervention. These mediators have been selected based on published evidence indicating that they are important in the relationship between health and work, and they are modifiable through interventions like vocational advice. The measures include personal (physical and mental health, and behaviours) and occupational measures and are summarised in Table 1.

### Electronic Medical Record (EMR) review

In addition to the data collection from the self-report questionnaires and SMS text messages, participants will have the option to consent for the research team to access their general practice EMR data for the study duration to allow examination of fit notes issued. Medical records for consenters will be extracted electronically and will be pseudonymised at source by the EMR system. Records will be exported by general practice staff with support from Clinical Research Network staff, as required.

## Data management

Data management will be carried out in accordance with the Study Data Management Plan designed by the TMG in accordance with Keele University Health and Social Care Research Quality Management System Standard Operating Procedures (HSCR Standard Operating Procedures (SOPs)). Questionnaires will be date stamped on receipt at the Keele CTU. Questionnaire data will be logged as returned on a management database and the participants’ responses entered / scanned into a database; the databases will be tested a priori for functionality and reliability. The study statistician will determine coding of questionnaire items, in accordance with standardised coding procedures as per relevant SOPs to facilitate data entry. Keele CTU staff will enter / scan data and data entry checks will be carried out as per relevant SOPs to ensure quality of data entry.

## Analysis

### Internal pilot

The internal pilot trial will assess recruitment and intervention fidelity over the first four months and be focused on the proportion with follow-up data at six weeks. The anticipated recruitment rate will be approximately four patients/practice/month in the internal pilot and aim to achieve 80% follow-up for the primary outcome at six months. We will scrutinise (i) recruitment (target versus actual, % eligible, % consenting and randomised), (ii) intervention fidelity, and (iii) follow-up rate at six weeks. A ‘stop (Red)/ amend (Amber)/ go (Green)’ set of progression criteria including: (i) recruitment uptake ≤70% of those eligible and consent to participate (Red), 71%-99% (Amber), 100% (Green); (ii) engagement with the intervention, % of intervention arm participants who have at least one contact with a VSW <40% (Red), 40-65% (Amber), >65% (Green); (iii) primary outcome data at six weeks follow-up rate <60% (Red), 60-80% (Amber), >80% (Green). A decision to continue to the main trial will be made if all progression criteria are met at the ‘Green’ level, and to continue but with some adjustments if any criteria are at least ‘Amber’; the trial may be stopped if any of the criteria are ‘Red’ and the Trial Management Group (TMG), Trial Steering Committee (TSC) and Data Monitoring Committee (DMC) agree they cannot be addressed.

### Main trial

Baseline participant characteristics will be summarised according to the nature of the data (mean/standard deviation for normally distributed variables; median/inter-quartile range for skewed numerical data; frequency/percent for categorical variables) – overall, and by treatment arm (no formal statistical testing will be carried out). An intention-to-treat analysis will be carried out as the main approach: analysing participants as per randomised allocation. This is in line with the pragmatic nature of the trial – allowing for infrequent referral to occupational health within both arms and a lack of contact with the vocational support intervention in a small proportion of participants in the intervention arm.

For the primary outcome measure of days off work, we will present descriptive statistics on both mean (and median) number of days off work with standard deviation (and interquartile range) – for the time intervals baseline to six weeks and six months follow-up, by trial arm. The inferential analysis will be carried out by negative binomial (or Poisson) regression models adjusting for age, sex, centre, main health condition for which the fit note was issued (MH, MSK, other) and time off work due to health condition in the months prior to trial participation (fixed effects). If there is over-dispersion (skewness) in the outcome data, then the negative binomial model will likely be preferred to the Poisson model; the goodness of fit of each model will be assessed to determine which model is most appropriate through scrutiny of the likelihood-ratio test and Akaike/Bayesian Information Criterion (AIC/BIC). The estimated effect will be presented as an incidence rate ratio; the results will be given as point estimates with 95% precision interval and associated p-value. The primary endpoint evaluation will be number of days off work over the six months follow-up, with days off work over the initial six weeks follow-up as a secondary endpoint.

Similarly, descriptive summaries of data (mean (SD) / median (IQR) / frequency count (percent)) by trial arm will be presented for secondary outcomes. Proportions of participants accessing each step of the intervention (1, 2 and 3) and the content of the intervention as detailed on CRFs will be reported for the intervention arm. A mixed-model approach will be carried out for between-arm estimation of mean differences (for numerical outcomes) or odds ratios (for categorical outcomes) through linear or logistic link functions, respectively. The regression models will include the same covariates as outlined above for the primary outcome analysis and additionally the corresponding baseline value (as appropriate) e.g. baseline RTW self-efficacy score for evaluation of between-arm difference in mean RTW self-efficacy at follow-up. Time to sustained RTW data, collected by fortnightly SMS text messages, will be evaluated through survival analysis methods: life table and Kaplan-Meier descriptive summaries and Cox regression modelling with covariates as detailed above.

A complier average causal effect (CACE) evaluation will be undertaken to obtain unbiased estimation for the comparison of the primary outcome for those participants in the intervention arm who had at least one contact with the VSW versus similar participants in the control arm. A small number of pre-specified subgroup analyses will be carried out evaluating whether between-arm differences in the primary outcome measure (number of days off work over six months) contrast across potential baseline moderators: baseline main health condition resulting in the fit note (MSK, MH, other) and duration of work absence in previous six months. Statistical estimates will be obtained through including interaction terms for trial arm × baseline subgroup within the statistical model of treatment effect. All statistical analyses are focused on superiority testing based on 5% two-tailed significance level.

Analyses will be carried out blind to intervention allocation (with the exception of the CACE analysis and CRF data analysis describing the content of the vocational advice intervention) and double-analysed by two statisticians. Data collection, checking and verification will be performed according to Keele CTU SOPs to ensure rigour. A detailed Statistical Analysis Plan (SAP) will be developed with guidance from the DMC and TSC to ensure transparency in the statistical analysis of the trial.

### Mediation analysis

Mediation analysis is a statistical approach for testing hypothesised causal pathways between variables thought to be important in explaining treatment outcome. Testing such causal mechanisms is an important aspect of process evaluation (35). Variables hypothesised to be key mediators of the intervention (reported in Table 1) will be finalised through the intervention adaptation in the feasibility phase. In the trial, data on these variables and on the primary outcome will be collected at baseline, 6 weeks and 6 months. Change in each mediator between baseline and each follow-up point will be described. Multilevel causal modelling techniques will be used to identify the proportion of the intervention effect on days absent from work explained by change in the potential mediators (indirect effects). Each mediator will be first analysed separately, then combined into a single multiple mediation model to assess the combined effect of the potential mediator variables on the outcome of days absent from work over six months. Latent growth curve models will be used for this analysis as they allow all three time points to be used within the analysis. These indirect effects on outcome will be expressed as a regression coefficient with bootstrapped 95% CI.

## Health economic evaluation

The health economic evaluation will be undertaken from the NHS and personal social service (PSS) perspective as recommended by the National Institute for Health and Care Excellence (NICE) (36). Secondary analyses will be undertaken from a healthcare and societal perspective. The analysis will follow standard recommended methods of health economic analyses and good practice guidance.

## Resource use and costs collection

Resource use and costs will be based on the standard approach used in economic evaluations following the three-stage process: identification of resource use, measurement, and valuation. Healthcare resource use will be collected using data from the six month postal questionnaires and general practice EMR reviews will describe issued fit notes. Healthcare costs will include primary and secondary care contacts such as GP and practice nurse consultations/home visits, medications, contacts with other healthcare professionals, NHS and private outpatient visits and inpatient stays and use of other vocational advice services. Information on patient-incurred costs will also be collected, such as over-the-counter purchases. Questions on time off work, presenteeism and occupation will provide information required to calculate the indirect (productivity) costs (benefits).

To calculate the cost of the vocational advice intervention, information on number and duration of contacts with the VSW (telephone calls, videoconference calls or face-to-face visits) will be obtained for each participant from the CRFs and unit costs applied. The costs of training and mentoring VSWs, and intervention delivery costs will also be calculated to inform decision-makers about total service costs and for inclusion in a sensitivity analysis.

Health resource use information obtained from the self-reported questionnaires at six months will be valued with unit cost data from standard sources, including the NHS reference costs (37), Unit Costs of Health and Social Care (38) and the British National Formulary (39). Due to the lack of nationally representative unit cost estimates for private healthcare, this care will be costed as the NHS equivalent.

## Health outcomes

The outcome measures for the cost-consequence analysis are self-reported number of days absent from work over six months, and health related quality of life measured by the responses to EQ-5D-5L questionnaire at baseline, six weeks and six months and benefits will be estimated from the data on productivity losses. The crosswalk value set will be applied to patient responses to obtain utility scores, in line with current NICE recommendations (40).

Productivity loss will be calculated using data collected on employment status at every time-point and number of days off work. Information on occupation, further details of typical work activities and the nature of their employment (full time or part time) will be requested. The average wage for each respondent will be identified using UK Standard Occupational Classification coding and annual earnings data for each job type (41). The analysis will use the human capital approach (42) and the self-reported days of absence will be multiplied by the respondent-specific wage rate. The human capital approach assumes that the value of lost work is equal to the number of resources an individual would have been paid to do that work, and values productivity losses because of morbidity (or mortality) by measuring time lost from work and multiplying this with the gross wage of the person. Whilst there is no standard tool for capturing the costs of presenteeism, we propose to use the Single-Item Presenteeism Question (SIPQ) contained within the Work Productivity and Activity Impairment Questionnaire (WPAI) (43). Our previous work has demonstrated this question to be valid and responsive in patients with MSK pain and other conditions (44). This estimation of perceived percentage loss in productivity can be applied to person-specific wage rates using the human capital approach. Given the many uncertainties in the measurement of costs due to presenteeism, this will be presented as part of a secondary analysis.

## Economic analysis (within trial-analysis)

Descriptive statistics will be presented for all costs and outcomes as means and standard deviations. Costs for the intervention and control arms will be presented and disaggregated for each of three cost categories (healthcare costs, patient-incurred costs, productivity costs). Total societal costs over the study period will be calculated by summing all items. Results on number of days absent from work will be obtained from the statistical team. Mean scores for the responses to EQ-5D-5L questionnaire at baseline, six weeks and six months will be presented by trial arm and will be combined with standard valuation sources to measure the Quality Adjusted Life-years (QALYs) gained. Differences in costs and QALYs will be described and an incremental cost-effectiveness ratio (ICER) calculated. The analysis will be based on imputed data, with adjustment for baseline covariates. Uncertainty around the base-case point estimates will be explored using bootstrapping on cost and QALY pairs to produce a cost-effectiveness plane and cost-effectiveness acceptability curve (CEAC).

A cost-benefit analysis will also be undertaken from a broad societal perspective to calculate the net societal benefit of the addition of the vocational advice intervention, by subtracting the difference in direct healthcare costs (costs) between the trial arms from the difference in indirect productivity costs (benefits) between the arms. This will also allow return on investment to be calculated by dividing the net benefits of the VSW intervention (gain minus cost) by the net costs of the intervention.

## Nested qualitative study

To explore experiences and perceived impact of the vocational advice intervention, a nested qualitative study will be undertaken with a sample of participants in the intervention arm.

## Methods

The questionnaire responses at 6 weeks will be screened to enable a purposive sampling frame to be applied to select participants for interview. A range of participant characteristics will be sampled for, including age, sex, health condition (MH, MSK, other condition), geographical area (West Midlands, London, Southampton), job type, duration of work absence, level of engagement with the vocational advice intervention (i.e., steps 1-3), and RTW status, with the aim of exploring the experiences of a broad range of participants about the vocational advice intervention and its impact on their progress to RTW.

Potential participants will also be informed when invited that if they agree to be interviewed, we will also ask for their permission to contact their primary care clinician, to invite them to agree to a separate, matched interview. We will also ask participants for permission to possibly contact their employer, line-manager, or supervisor (depending on who is most appropriate), to invite them for an interview. Only those employers/line-managers who have been contacted by the VSW as part of step 3 of the intervention will be invited for an interview, allowing us to explore their experiences of engaging with the intervention. This means that it is possible that only a small number of employers/line-managers will be invited for an interview.

These matched interviews will discuss the participant’s case, as well as accessing clinician and employer views on managing sickness absence more broadly. Potential participants will, however, be informed that they can still take part in an interview even if they do not agree to us contacting their clinician or employer. Those participants who consent to this at the time of their interview will be asked to provide the relevant contact details. Following the interview with the participant, the clinician will be contacted either by telephone, email, or post, following which an invitation letter and information sheet about the study will be emailed or posted to them.

Semi-structured interviews with up to 20 trial participants from the intervention arm, and up to 20 VSWs, healthcare professionals and employers/line-managers who have patients/employees in the intervention arm will be undertaken. The final number of interviews will be guided by data saturation, defined in terms of ‘informational redundancy’ =:J the point at which additional data no longer offers new insights (45). We anticipate all interviews will be conducted by telephone, but in-person interviews will be available if necessary. Topic guides will be used, informed by the study objectives and focused on understanding how the intervention works using the study specific logic model. Separate topic guides will be developed for trial participants, VSWs, healthcare professionals and employers’/ line-managers’ interviews. The topic guides will be used to prompt participants about a range of aspects relating to their experience of work absence including (but not limited to) the following:

- Perspectives and experiences of trial participants, VSWs, healthcare professionals, and employers/line-managers regarding work absence
- The influences on each groups’ decision-making around sickness absence
- The VSWs’ experiences of delivering the intervention; in particular, how they supported participants through the RTW process including their decision-making about the steps of intervention delivered (i.e., steps 1-3)
- The participants’ experiences of engagement in the vocational advice intervention, acceptability, mode of delivery, and whether/how it supported their RTW
- Participants’ decision-making in deciding to RTW
- Primary care clinicians’ views about the impact of the intervention on their own practice, e.g., number of fit notes issued, patients re-consulting, discussions with patients about work absence
- Impact of COVID-19 on the suitability of the WAVE intervention.

### Analysis

All interviews will be audio-recorded, fully transcribed and anonymised. An inductive, exploratory framework will be adopted using thematic analysis, and influenced by grounded theory(46). First, a sample of early transcripts will be independently coded by two researchers with experience in qualitative analysis, and a coding framework agreed upon, which will be applied in subsequent coding. Coded data will be analysed by the qualitative social science researcher and a second research team member independently to develop categories and themes for discussion. The constant comparison method (47) will be used in the analysis, looking for connections within and across interviews, and across codes, highlighting data consistencies and variation. Analysis will be an iterative process, with emergent findings used to further refine topic guides for subsequent interviews. Comparisons will then be made using a framework approach (48) between the experiences of participants with different health conditions looking for similarities and differences in the separate accounts particularly related to how the methods of identification, recruitment and intervention delivery can be improved. We will also use the acceptability framework of Sekhon *et al* (49) to sensitise the analysis.

## Discussion

The WAVE trial is testing the clinical and cost-effectiveness of the additional offer of a vocational advice intervention to usual primary care for adults receiving a fit note for time off work for mental health, musculoskeletal or other health conditions. Given the limited access to vocational healthcare in the UK and the need to access fit notes from primary care, we have developed and are testing an intervention delivered remotely by trained vocational support workers to address the issues of health and work early in patients’ work absence. The results of the WAVE trial will inform primary care practices and may guide the development of services to support people with health conditions and work issues. The primary outcome is number of days off work over six months, a range of secondary outcomes will also be assessed, and qualitative interviews will explore the value of the vocational advice intervention to patients, primary care clinicians, employers and vocational support workers.

## Data Availability

This is a protocol for a trial and no data is available from the current manuscript. D
All data produced as part of the trial will be available on reasonable request to the authors and following Keele University Data Request procedures.

